# Prevalence of Mild Cognitive Impairment in Mexican Older Adults: Data from the Mexican Health and Aging Study (MHAS)

**DOI:** 10.1101/2020.01.03.20016345

**Authors:** Miguel Arce Rentería, Jennifer J. Manly, Jet M. J. Vonk, Silvia Mejia Arango, Alejandra Michaels Obregon, Rafael Samper-Ternent, Rebeca Wong, Richard Mayeux, Sandra Barral, Giuseppe Tosto

**Author notes:** **Corresponding Authors:** Sandra Barral, Ph.D., Giuseppe Tosto, M.D., Department of Neurology, Taub Institute for Research on Alzheimer’s Disease and the Aging Brain, Columbia University Medical Center, 622 W 168th St, New York, NY, 10032. Last authors have contributed equally (Sandra Barral, Giuseppe Tosto).

## Abstract

**INTRODUCTION:** We estimated the prevalence and risk factors for mild cognitive impairment (MCI) and its subtypes in Mexican population using the cognitive aging ancillary study of the Mexican Health and Aging Study.

**METHODS:** Using a robust norms approach and comprehensive neuropsychological criteria, we determined MCI in a sample of adult Mexicans (N=1,807;55-97years). Additionally, we determined prevalence rates using traditional criteria.

**RESULTS:** Prevalence of amnestic MCI was 5.9%. Other MCI subtypes ranged 4.3% to 7.7%. MCI with and without memory impairment was associated with older age and rurality. Depression, diabetes and low educational attainment were associated with MCI without memory impairment. Using traditional criteria, prevalence of MCI was lower (2.2% amnestic MCI, other subtypes ranged 1.3%-2.4%).

**DISCUSSION:** Older age, depression, low education, diabetes, and rurality were associated with increased risk of MCI among older adults in Mexico. Our findings suggest that the causes of cognitive impairment are likely multifactorial and may vary by MCI subtype.

**Research in Context:** *Systematic review:* We reviewed the literature using Google Scholar and PubMed. Few studies have reported prevalence rates for mild cognitive impairment (MCI) in Mexican population. These studies have primarily relied on limited cognitive assessments, and diverse MCI criteria. Evaluating the prevalence of MCI with a robust neuropsychological approach can help understand the rates and risk factors associated with MCI across a large and representative sample of the aging Mexican population.

*Interpretation:* Various sociodemographic and health factors such as older age, depression, low education, diabetes, and rurality were significant correlates of MCI and differed by MCI subtype.

*Future directions:* Longitudinal studies will be needed to evaluate the diagnostic stability of MCI over time, and its association with incident dementia. Future work will evaluate the casual path of these sociodemographic and health factors on cognitive impairment to develop effective interventions.

## Background

By 2050, two-thirds of older adults with dementia will live in low-and middle-income countries (LMIC).^1^ As these LMIC continue to experience a reduction in mortality rates and improved access to healthcare, it is critical to understand the factors that can lead to increased dementia risk.^2,3^ Mild cognitive impairment (MCI) is a diagnostic entity associated with increased risk of incident dementia.^4^ The most widely used approach to diagnose MCI across various clinical trials^5^ and population-based studies^6^ relies on simple cognitive screening measures, limited neuropsychological assessment that generally uses one test to define a cognitive domain, and the inclusion of self-reported memory complaints. Using neuropsychological approaches to classify MCI, that uses a full range of neuropsychological measures with actuarial decision-making, has been associated with greater diagnostic stability overtime (e.g., less likelihood of reversal to cognitively normal status), reduced false-positive diagnoses, incident dementia, and with Alzheimer’s disease (AD) biomarkers compared to conventional “one test” diagnostic approaches.^7,8^ Evaluating the prevalence of MCI with a robust neuropsychological approach can help identify individuals at greater risk for dementia and improve our understanding of the potential sociodemographic and health factors associated with MCI in LMIC.

Recent studies have used different diagnostic criteria to estimate prevalence rates for MCI in the Mexican population. Our group reported a 25% prevalence rate for cognitive impairment with no dementia (CIND) relying on a brief cognitive screening test (the Cross-Cultural Cognitive Examination, CCCE).^9,10^ However, this approach may lead to diagnostic misclassification, i.e., increase rates of false-positive and false-negatives. Similarly the 10/66 Dementia Research Group,^11^ relied on a brief cognitive assessment supplemented with additional memory tests and reported a prevalence rate of 3.2% for the amnestic subtype of MCI^6^ only. Using a comprehensive neuropsychological evaluation,^11^ Juarez-Cedillo and colleagues estimated MCI prevalence rates at 6.45% among primary insurance beneficiaries in Mexico City. To classify participants as MCI, the study relied on a single test per cognitive domain. However, employing multiple tests per cognitive domain to define impairment provides a more reliable estimate.^12^ Moreover, the study’s sample was not representative of the general Mexican population as they were primarily adults living in Mexico City with access to government and employer co-sponsored healthcare. To understand the rates and risk factors associated with MCI across the aging Mexican population, a large and representative sample of older adults is needed.

The Mexican Health and Aging Study (MHAS) started in 2001, with a nationally representative sample of adults over the age of 50 in both urban and rural areas of Mexico designed to prospectively evaluate the impact of disease on health, function and mortality.^13^ The study protocols and survey instruments are highly comparable to the U.S. Health and Retirement Study.^14^ In 2016, MHAS launched an Ancillary Study on Cognitive Aging in Mexico (Mex-Cog) among a sub-sample of MHAS participants. The goal of Mex-Cog was to perform a comprehensive cognitive assessment using the same harmonized cognitive assessment protocol currently used by other ongoing population-based longitudinal studies of aging around the world.^15^ The Mex-Cog study provides a unique opportunity to estimate MCI in a large and representative sample of Mexican adults.

The primary goals of the current study are i) to define diagnostic criteria for MCI in an elderly Mexican population using an actuarial neuropsychological approach, ii) to establish MCI prevalence rates, and iii) to evaluate the association between MCI and sociodemographic and health factors.

## Methods

### Study Participants

Participants were a sub-sample of the MHAS 2015 wave who received the Mex-Cog cognitive battery. Full study procedures and descriptions for MHAS have previously been reported.^13^ The study was approved by the Institutional Review Boards of the University of Texas Medical Branch in the United States and the National Institute of Public Health in Mexico.

MHAS participants, 55 years and older, were selected from eight different states for geographic convenience and characterized by a balanced proportion of urban and rural population, high prevalence of diabetes, high migration history to the United States, high presence of mining or metallurgical industry, and high presence of pottery industry. Overall, 2,265 MHAS participants completed the in-person household interview and 2,042 participants were administered the Mex-Cog assessment.

As described in the Mex-Cog protocols, participants complete a short or long version of the cognitive assessment based on their performance on a modified version of the Mini-Mental State Examination (MMSE).^16^ Participants who had a MMSE score ≤10 (n=102) were considered highly impaired and were not administered the full Mex-Cog battery and thus were not included in the current sample. Additional exclusion criteria included missing demographic (n=17) and/or neuropsychological data (n=23) and participants classified as dementia cases based on the MHAS 2015 criteria (n=93). The final analysis sample included 1,807 MHAS participants (89% of the initial sample). Figure 1 shows the flow chart for the sample’s selection criteria.

**Figure 1.**
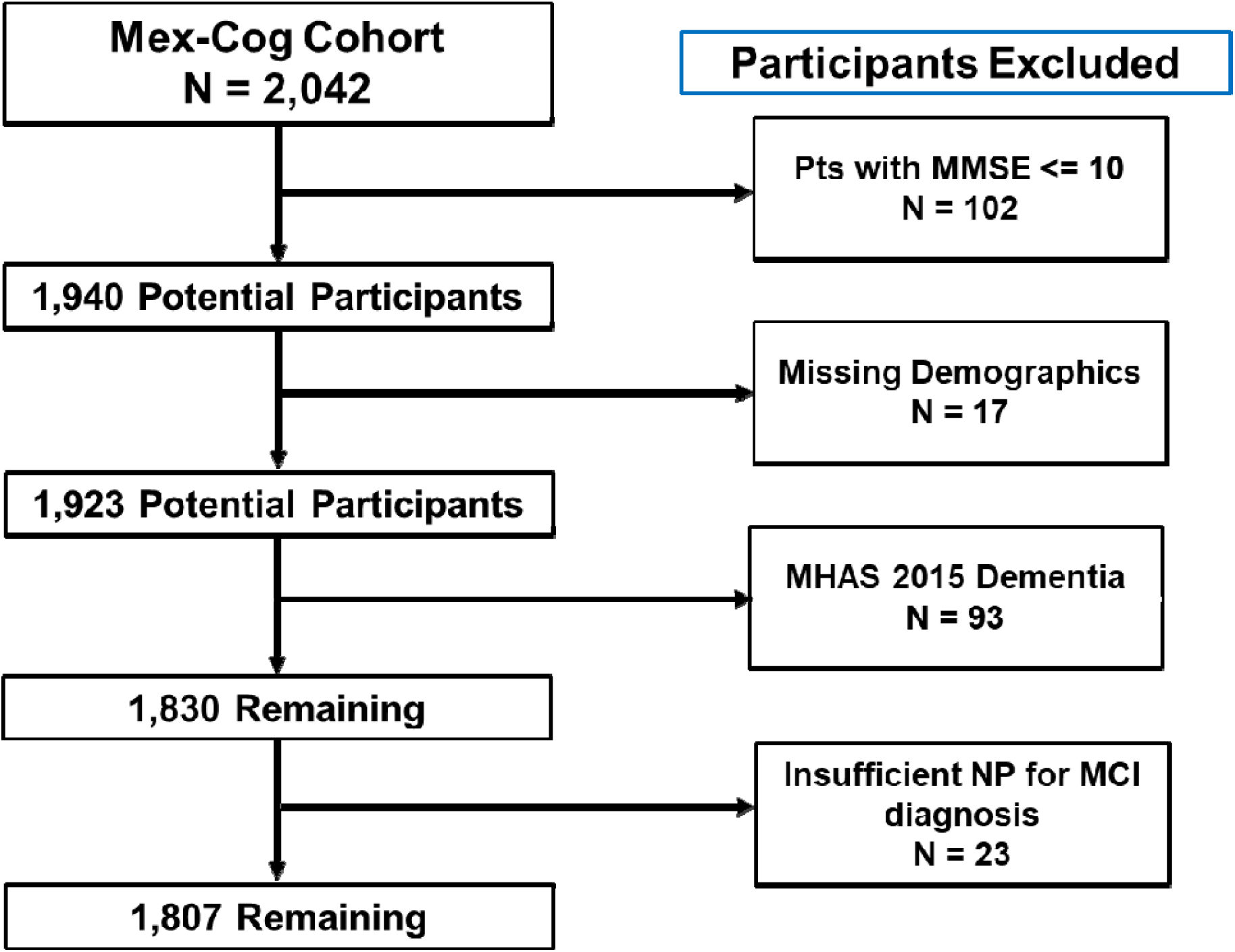
Flow diagram for identifying participants for the current sample.

### Cognitive Assessment

Participants completed a 28-point modified version of the MMSE^17^ and a comprehensive cognitive assessment evaluating various cognitive domains such as memory, language, visuospatial function, and executive functioning. Supplementary Table 1 describes the cognitive battery by domain. All test scores were standardized into z-scores using the mean and standard deviation from the entire sample. Within each cognitive domain, composite scores for each domain were calculated by averaging the z-scores.

### Self-Report Assessment

In addition to the cognitive assessment, participants answer questions regarding memory complaints and depressive symptoms using a modified version of the Center for Epidemiologic Studies Depression Scale (CESD).^18^

### Informant Interview

The Mex-Cog protocol included an interview with an informant: a person that was familiar with the behavior and health of the participant, most frequently a spouse, an adult child, or a caregiver. The Mex-Cog informant instrument included 28-items from the Community Screening Interview for Dementia (CSID)^19^ that asks about the participant’s performance in everyday living, and an adapted version of the History and Aethiology Scale^20^ to assess history of cognitive decline. Greater detail on the informant interview can be found on the Mex-Cog methodological document.^16^

### Sociodemographic and Health Factors

The MHAS 2015 visit collected data on rurality, i.e., whether the participant resides in urban or rural areas based on community population density cut points using standard values used by the Mexican statistical bureau (Instituto Nacional de Estadística y Geografía; INEGI). Rurality was categorized into four levels: 1)100,000+ residents, 2)between 15,000-99,999 residents, 3)between 2,500-14,999 residents, and 4)<2,500 residents. For the current study, we dichotomized rurality as participants living in an urban area (≥100,000 population size) compared to all other groups.

A cardiovascular disease burden score was derived by combining the number of cardiovascular diseases endorsed (hypertension, diabetes, heart disease, and stroke). Current workforce status was ascertained by self-report as either unemployed, retired, or employed. Health care availability was defined as whether participants have access or not to health insurance (governmental or private).

### Regression-Based Neuropsychological Test Norms

The normative sample used to define MCI was selected by means of a robust norms approach. An ideal robust norms approach would include participants that do not develop dementia over time. However, due to the lack of follow-up cognitive data for Mex-Cog participants, to infer absence of clinically significant cognitive decline, we used the informant report. The normative sample excluded participants who met criteria for impairment in their MHAS 2015 visit (i.e., CIND, dementia), reported stroke, severe depressive symptoms and those with significant cognitive decline according to the informant’s report. Significant cognitive decline was operationalized as the informant endorsing any of the following items on the CSID: 1) regularly forgets names of family members; 2) regularly uses wrong words; 3) regularly forgets when they last saw informant; 4) forgets what happened the day before; 5) forgets where he/she is; 6) gets lost in their own neighborhood; 7) gets lost at home; 8) change in the ability to think and reason; 9) mistook a family member with another person; or 10) reasoning is confusing or illogical. We also excluded participants whose informant endorsed functional decline that could likely be attributable to significant cognitive decline or depression such as endorsing either 1) stopped doing activities or hobbies or 2) change in the ability to handle money. A total of 547 participants were selected as the normative sample (Supplementary Table 2).

A regression-based approach was used to develop demographically corrected T-scores.^21,22^ Multiple linear regression analyses were run to evaluate the influence of age, gender, and years of education on each of the cognitive domains using the normative sample. We then used the resulting beta coefficients and standard error of each regression model to calculate predicted scores for each cognitive composite (i.e., expected scores based on the participant’s age, gender, and education) across the entire sample (N= 1,830). A residual score was calculated by then subtracting each participant’s predicted composite score from their actual composite score. Lastly, residual scores were converted to T-scores according to the following formula: T score= [(Residual Score/SE of Estimate for the Regression Equation)*10] + 50.

### MCI Neuropsychological Classification

Participants were classified as MCI following an actuarial neuropsychological test approach.^7,23^ Participants were considered MCI if a composite cognitive domain score was ≥ 1.5 SD below demographically-corrected T-scores (participants with a cognitive domain T-score ≤ 35). We then characterized the MCI group by subtype as either single-domain or multiple-domain MCI based on the type and number of cognitive domains impaired (i.e., MCI-amnestic, MCI-language, MCI-executive function, MCI-visuospatial, multiple domain amnestic, multiple domain non-amnestic). Lastly, given that prior studies indicate that MCI with memory impairment has a stronger association with AD biomarkers and greater likelihood to decline over time than MCI without memory impairment,^7^ we dichotomized our MCI groups as either with or without memory impairment, regardless of it being single- or multiple-domain as done in previous studies.^22,24^

### Statistical Analyses

MCI prevalence rates and 95% confidence intervals (95%CI) were calculated for the overall sample, and stratified by age (median split 66), years of education (median split 6), and gender. We assumed a Poisson distribution when determining the 95%CIfor the prevalence rates. Chi-square tests evaluated the relationship of each MCI subtype by age, years of education, and gender. Descriptive statistics were conducted in SPSS 26.^25^ Multinomial logistic regressions to evaluate the association between MCI and sociodemographic and health factors were conducted in R Studio.^26^ We additionally determined a modified MCI diagnostic criteria that incorporated self-reported memory complaints (“do you feel your memory is worse than before?”) in order to present prevalence rates using traditional criteria.^4^

## Results

Table 1 describes the overall sample’s characteristics. Fifty-nine percent of the sample were women, with an average age of 67 ±8 years, and mean education level of 6 years ± 5. The majority of the participants (58%) lived in urban area (population size 100,000+). Depressive symptoms above the modified CESD cut-off was reported in 36% of the sample. As to disease burden, around 60% of the sample endorsed being diagnosed with one or more cardiovascular conditions. Almost half of the participants (47%) reported hypertension, while the prevalence of diabetes (27%), heart disease (9%), and stroke (2%) was lower across the sample. The normative sample was on average 66 years of age (SD=7.8, range 54-97 years), with an average of 7 years of education (SD=4.9, range 0-19 years), and 59% women.

**Table 1.** Total Sample Characteristics and by MCI Diagnosis (*N* = 1,807)

| **Table 1. Total Sample Characteristics and by MCI Diagnosis (N = 1,807)** |  |  |  |  |  |
| --- | --- | --- | --- | --- | --- |
|  | All<br>(N = 1,807) | non-MCI<br>(n = 1,185) | MCI with<br>Memory<br>Impairment<br>(n = 235) | MCI without<br>Memory<br>Impairment<br>(n = 387) | *p-value* |
| Age; *avg(SD)* | 67.17 (8.37) | 66.40 (7.87) | 68.60 (9.70) | 68.68 (8.67) | <0.001 |
| Education; *avg(SD)* | 5.67 (4.48) | 6.20 (4.31) | 5.43 (4.65) | 4.17 (4.52) | <0.001 |
| Gender |  |  |  |  |  |
| % women | 58.9 | 57.6 | 63.8 | 59.9 | .181 |
| Depressive<br>Symptoms |  |  |  |  |  |
| CESD; *avg(SD)* | 3.71 (2.68) | 3.50 (2.69) | 3.92 (2.54) | 4.25 (2.67) | <0.001 |
| Rurality; % |  |  |  |  | <0.001 |
| 100,000 + | 57.9 | 61.6 | 52.8 | 49.6 |  |
| 99,999-15,000 | 14.7 | 14.9 | 11.1 | 16.3 |  |
| 14,999-2,500 | 8.4 | 7.6 | 10.2 | 9.8 |  |
| <2,500 | 19.0 | 15.9 | 26.0 | 24.3 |  |
| % Insured | 91.0 | 91.3 | 91.1 | 90.2 | .802 |
| Employment Status |  |  |  |  | .001 |
| % Employed | 35.7 | 37.1 | 27.8 | 36.4 |  |
| % Retired | 16.2 | 17.9 | 14.5 | 11.9 |  |
| % Unemployed | 48.1 | 45.0 | 57.7 | 51.7 |  |
| Medical Conditions |  |  |  |  |  |
| Disease Burden<br>Score; *avg(SD)* | 0.88 (0.91) | 0.87 (0.92) | 0.79 (0.81) | 0.93 (0.93) | .183 |
| Hypertension; % | 47.3 | 47.4 | 46.6 | 47.4 | .972 |
| Diabetes; % | 26.6 | 25.5 | 24.8 | 31.3 | .065 |
| Heart Disease; % | 9.3 | 10.2 | 6.0 | 8.7 | .117 |
| Stroke; % | 2.3 | 1.9 | 1.7 | 3.6 | .130 |
| CESD = Center for Epidemiologic Studies Depression Scale; NS= non-significant, p>0.05. |  |  |  |  |  |

### Frequency of MCI

Across 1,807 participants, 13% met criteria for MCI with memory impairment and 21% for MCI without memory impairment (total=34%). Table 1 also describes the characteristics of MCI with and without memory impairment. Compared to non-MCI individuals, MCI participants with and without memory impairment were an average of three years older, had fewer years of education, endorsed higher prevalence of depressive symptoms, and were more likely to live in a rural setting. There was a greater proportion of participants unemployed among those with MCI with (58%) and without memory impairment (52%) compared to the non-MCI group (45%).

Table 2 describes the prevalence rate for overall and subtypes of MCI by age, education, and sex. Across the entire sample, the prevalence rates of MCI subtypes ranged from 4.2% with isolated impairment in executive functioning to 7.7% with isolated visuospatial impairment. The frequency of overall MCI was higher among those older than 66 years of age (*p*<0.001) and less education (*p*<0.001). As for MCI subtypes, participants that were older were more likely to have isolated deficits in visuospatial abilities (*p*=0.043), and multiple domain amnestic MCI (*p*=0.033). Less educated participants were more likely to have isolated deficits in language (*p*=0.004), visuospatial abilities (*p*<0.001), executive functioning (*p*=0.007), multiple domain amnestic MCI (*p*<0.001) and non-amnestic multiple domain MCI (*p*<0.001). Women were more likely to have non-amnestic multiple domain MCI than men (*p*=0.023).

**Table 2.**
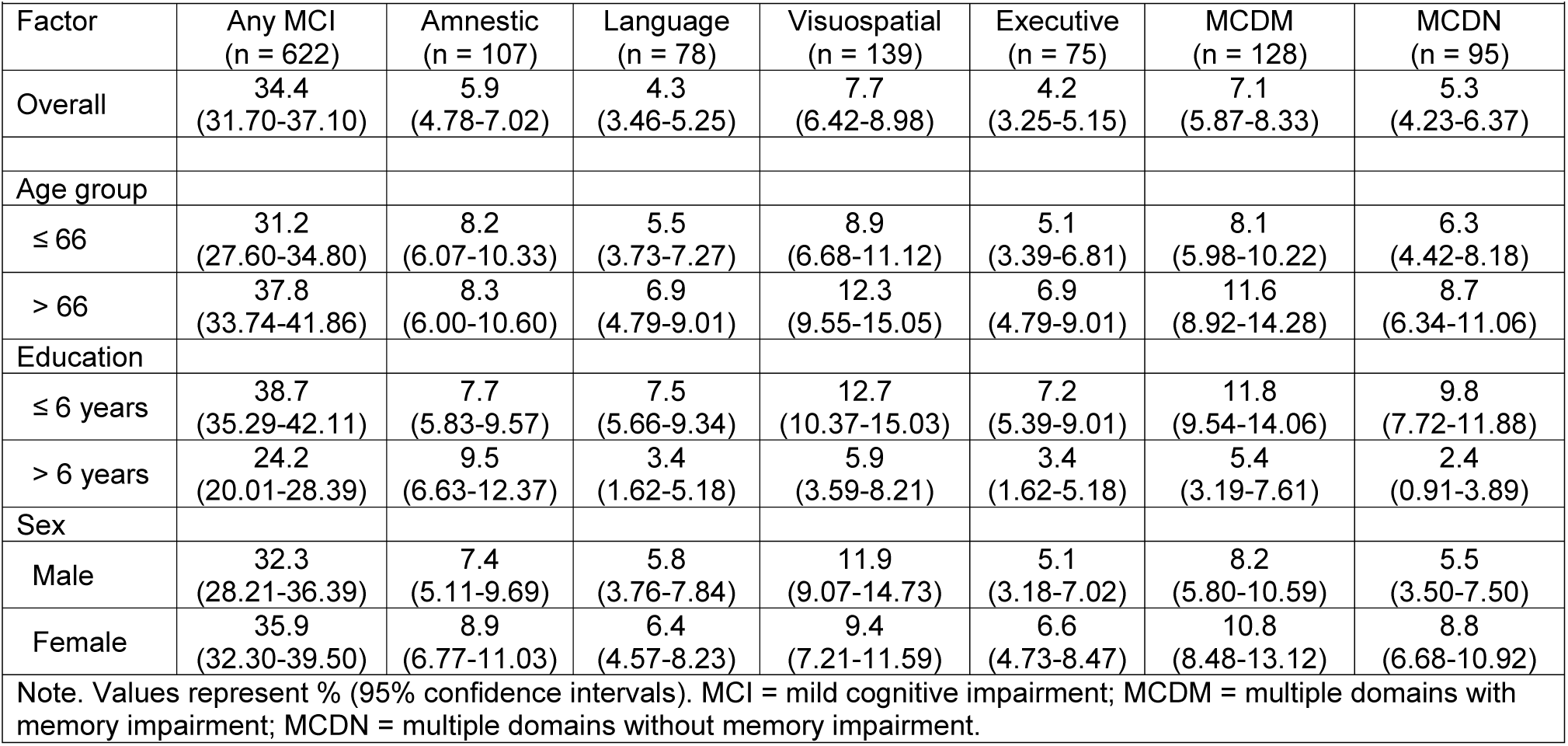
Prevalence of MCI.

Supplementary Table 3 provides prevalence rates for participants who met criteria for MCI that incorporated memory complaints in the operational criteria (overall %=???). As expected, the overall rate of MCI decreased by more than half with rates ranging from 1.3% for isolated deficits in language to 3.0% for isolated impairment in visuospatial abilities. However, the relationship between age, education, and gender with MCI subtypes did not differ from that previously reported. Older age was associated with isolated impairments in visuospatial abilities (*p*=0.01) and multiple domain-amnestic MCI (*p*=0.04). Fewer years of education was more likely to be associated with isolated impairments in visuospatial abilities (*p*=0.01), and multiple domain non-amnestic MCI (*p*=0.02). Lastly, women were more likely to have single domain amnestic MCI (*p=*0.018) and non-amnestic multiple domain MCI (*p*=0.021).

### Risk Factors for MCI

Results of the multinomial logistic regression (Table 3) showed that older age and residing in a rural setting were associated with increased risk of MCI with memory impairment. Greater risk of MCI without memory impairment was associated with older age, less education, higher prevalence of depressive symptoms, residing in a rural setting and history of diabetes. Being retired compared to employed was associated with a reduced risk of MCI without memory impairment. Prior history of heart disease was associated counterintuitively with a reduced risk of MCI with and without memory impairment. Sex, stroke, hypertension, and insurance were not associated with increased risk of MCI with or without memory impairment (*p*>0.05).

**Table 3.** Multinomial Logistic Regression Predicting MCI Status.

| Table 3. Multinomial Logistic Regression Predicting MCI Status |  |  |  |  |
| --- | --- | --- | --- | --- |
|  | B (SE) | OR | 95% CI for OR |  |
|  |  |  | Lower | Upper |
| <u>MCI-Memory vs Unimpaired</u> |  |  |  |  |
| Age | 0.03 (.01) | 1.03** | 1.01 | 1.05 |
| Education | 0.02 (.02) | 1.02 | 0.98 | 1.06 |
| Sex | 0.19 (.18) | 1.21 | 0.85 | 1.74 |
| Rurality | 0.40 (.16) | 1.49* | 1.08 | 2.06 |
| Depressive Symptoms | 0.05 (.03) | 1.05 | 0.99 | 1.12 |
| Medical Conditions |  |  |  |  |
| Stroke | 0.08 (.56) | 1.08 | 0.36 | 3.21 |
| Hypertension | -0.10 (.16) | 0.91 | 0.66 | 1.24 |
| Diabetes | -0.07 (.18) | 0.93 | 0.65 | 1.33 |
| Heart Disease | -0.62 (.31) | 0.54* | 0.29 | 0.99 |
| Insurance Status | 0.08 (.26) | 1.08 | 0.65 | 1.82 |
| Employment Status |  |  |  |  |
| Retired | -0.03 (.26) | 0.97 | 0.59 | 1.61 |
| Unemployed | -0.14 (.20) | 1.33 | 0.90 | 1.96 |
| <u>MCI-Non-Memory vs Unimpaired</u> |  |  |  |  |
| Age | 0.02 (.01) | 1.03** | 1.01 | 1.04 |
| Education | -0.06 (.02) | 0.94*** | 0.91 | 0.97 |
| Sex | 0.01 (.15) | 1.01 | 0.75 | 1.36 |
| Rurality | 0.30 (.14) | 1.35* | 1.03 | 1.77 |
| Depressive Symptoms | 0.07 (.03) | 1.07** | 1.02 | 1.12 |
| Medical Conditions |  |  |  |  |
| Stroke | 0.61 (.38) | 1.84 | 0.88 | 3.85 |
| Hypertension | -0.15 (.14) | 0.86 | 0.66 | 1.12 |
| Diabetes | 0.31 (.14) | 1.37* | 1.03 | 1.82 |
| Heart Disease | -0.49 (.23) | 0.61* | 0.38 | 0.98 |
| Insurance Status | -0.07 (.23) | 0.93 | 0.60 | 1.45 |
| Employment Status |  |  |  |  |
| Retired | -0.47 (.22) | 0.63* | 0.40 | 0.97 |
| Unemployed | -0.14 (.16) | 0.87 | 0.64 | 1.19 |
| *Note.* Age and education were mean centered, male sex, urban setting, absence of medical condition, being insured, and employed were the reference groups; ***<br>$p<.001$, **$p<.01$, *$p<.05$; MCI=mild cognitive impairment. | | | | |

Supplementary Table 4 displays the results of a multinomial logistic regression within participants who met criteria for MCI with memory complaints. Results remained largely unchanged other than rurality, heart disease, and workforce status were no longer associated with risk of MCI.

## Discussion

MCI is a risk factor for dementia,^4^ as such, accurate ascertainment of the prevalence of MCI is critical to understand cognitive health of the population and factors associated with risk of MCI. Using data from a comprehensive cognitive assessment in a large cohort of older adults in Mexico (Mex-Cog), prevalence of MCI in this population was estimated as 13% and 21% for MCI with and without memory impairment, respectively.

Prevalence rates from international population based studies report great variability on the rates of MCI with some studies reporting rates for any MCI ranging from 3% to 42%, and amnestic MCI between 0.5% to 31.9%.^27^ In studies of Mexican population, the estimates of MCI prevalence are also highly variable, most likely due to the differences in MCI criteria and characteristics of the specific samples evaluated. Previous work with the 2001 wave of MHAS reported prevalence rates for CIND of 25%.^10^ Several methodological differences may explain the discrepancy in rates of MCI between these two MHAS studies. In the 2001 MHAS study^10^ the criteria for MCI was based on a brief dementia screening instrument (CCCE), while for the Mex-Cog study we used a comprehensive cognitive assessment. Relying on a cognitive screener may lead to a greater overestimation of cognitive impairment because of its coarse measurement and inability to assess impairment per cognitive domain.^21,28,29^ The 10/66 Dementia Research Group reported a prevalence rate of 3.2% for amnestic MCI among a diverse sample of older Mexicans.^6^ While our study provided a prevalence rate slightly higher (5.9%) for amnestic MCI, a key methodological difference was that the 10/66 research group relied on the Petersen criteria^30^ which incorporates memory complaints as part of the MCI diagnostic criteria. When memory complaints are included in our MCI diagnostic criteria, the prevalence of amnestic MCI in our sample? was 2.2%. Juarez-Cedillo et al. reported prevalence of MCI among older healthcare beneficiaries residing in Mexico City of 0.3% for multiple domain non-amnestic MCI and 2.6% for multiple domain amnestic MCI.^31^ While participants in the Juarez-Cedillo study^31^ were evaluated with a comprehensive cognitive assessment, it was heavily weighted on assessing memory, it was unclear which other domains were evaluated, and included memory complaints in their diagnostic criteria. In addition, most of the normative standards for their cognitive instruments were derived from non-Mexican Spanish-speaking populations (i.e., the Syndrom Kurztest used in their study was validated among elderly Chileans)^32^ which can limit the reliability to detect cognitive impairment. Lastly, all participants in the Juarez-Cedillo et al. study^31^ were residing in Mexico City and had access to healthcare through the Mexican Social Security Institute (IMSS) which provides health insurance to workers in the formal labor market. In contrast, almost half (42%) of the participants in the current study resided in less populated settings and only 54% reported receiving their healthcare through the IMSS.

As previously reported, older age, fewer years of education, and greater depressive symptoms increased the risk of MCI. Studies of Mexican population have reported on the association between age, education, depression and cognitive impairment.^33,10,31^ Results evaluating the impact of gender on the risk of MCI have been conflicting. Some studies reported higher risk of MCI among women,^10,31^ others among men,^34^ while others found no gender differences.^35^ In the current study, we did not find gender differences in risk of MCI. Additional sociodemographic factors such as rurality were associated with greater risk of MCI. Prior work has associated poorer cognitive functioning among older Mexican adults residing in more rural areas.^36^ The association of rurality and cognition may be in part driven by the historical educational disadvantage found in rural areas.^36^ In our current study, both years of education and rurality contributed independently to the increased risk of MCI. Medical conditions such as diabetes and heart disease were associated with MCI risk. While diabetes increased risk of MCI, a finding that is in line with prior studies among older Mexican adults,^37,38^ heart failure was associated with a reduced risk of cognitive impairment. Studies examining the association between heart disease and cognitive impairment have yielded inconsistent results. A potential reason is the complexity of assessing the entire spectrum of heart disease such that frequently only heart disease severe enough to result in a cardiac event such as a myocardial infarction is investigated.^39-41^ Consequently, subjects with severe heart disease who are not surgical candidates, and those with less severe heart disease (e.g., stable angina), may not be included in studies assessing the association of heart disease with cognitive impairment.^42^ In addition, since study participants with heart disease were more likely to be insured (7.9% versus 2.5%, *p*=0.010), it is possible that non-MCI participants are more likely to have greater access and utilization of healthcare services,^43^ and therefore show higher prevalence of diagnosed heart disease. Moreover, the self-reported nature of heart disease may contribute to spurious associations. Finally, our results showed that retirees were at lower risk of MCI compared to those employed. When compared to the unemployment group, the retired group appeared to be higher educated (average years of education of 8 versus 6 respectively, p<0.001). It is possible that higher educational attainment and other socioeconomic factors contribute to the lower risk of MCI among retirees.

A strength of the study was the use of an actuarial neuropsychological approach to defining MCI. Different MCI subtypes are associated with unique brain-behavior traits. For instance, amnestic MCI is associated with greater cortical thinning of temporal structures,^44^ while MCI with deficits in executive functioning is associated with white matter lesions.^45^ Similarly, MCI diagnoses derived with a neuropsychological approach when compared to MCI classifications using traditional criteria (i.e., Petersen et al.)^30^ were more likely over time to remain as MCI or progress to dementia, less likely to be reclassified as cognitively normal, more likely to be APOE-4 carriers and demonstrate abnormal cerebrospinal fluid AD-biomarker levels.^7^ Future studies including biomarkers will be needed to evaluate the diagnostic stability of MCI over time.

Several limitations deserve mention. First, further work is warranted to further characterize the current cognitive assessment and determine its measurement invariance by examining whether it measures the same cognitive constructs across different subpopulations (e.g., educational gradients).^46^ Second, due to the lack of follow up data for Mex-Cog participants, we relied on informant report to determine both clinically significant cognitive and functional decline.^47^ Third, due to the lack of a clinical diagnosis of dementia or detailed information on instrumental activities of daily living at the Mex-Cog visit, participants classified as MCI may very well meet criteria for clinical diagnosis of dementia.

Understanding the prevalence and factors associated with MCI can help elucidate the determinants of MCI and subsequent dementia in order to inform research and policy for preventative strategies. Establishing a protocol to define MCI using a neuropsychological approach across large studies of health and aging may improve our understanding of cognitive health among low-and middle-income countries such as Mexico.

## Data Availability

data available upon request at http://www.mhasweb.org/Data.aspx

---

**Supplementary Table 1.**
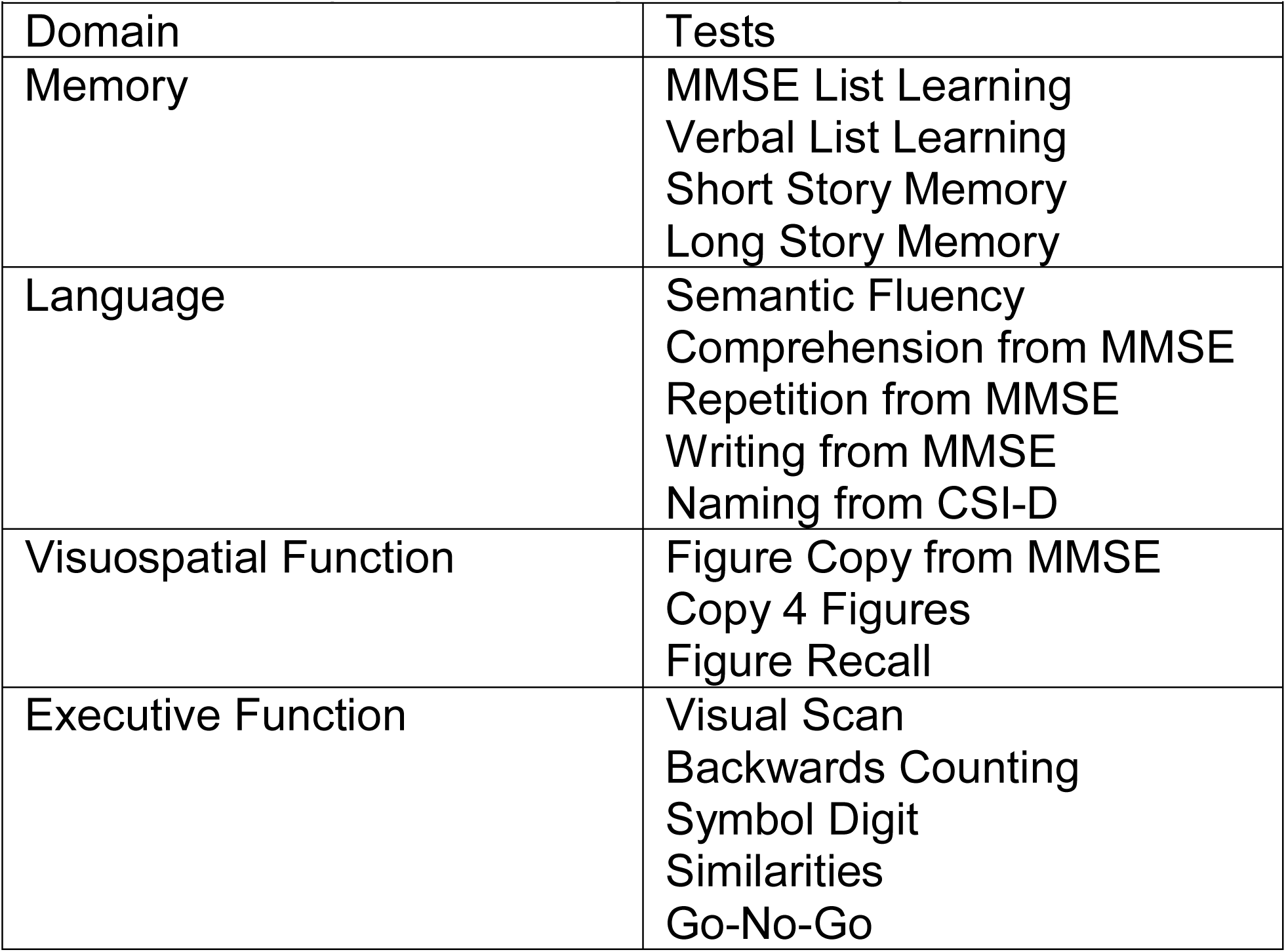
Cognitive Battery.

| **Supplementary Table 1. Cognitive Battery** |  |
| --- | --- |
| Domain | Tests |
| Memory | MMSE List Learning<br>Verbal List Learning<br>Short Story Memory<br>Long Story Memory |
| Language | Semantic Fluency<br>Comprehension from MMSE<br>Repetition from MMSE<br>Writing from MMSE<br>Naming from CSI-D |
| Visuospatial Function | Figure Copy from MMSE<br>Copy 4 Figures<br>Figure Recall |
| Executive Function | Visual Scan<br>Backwards Counting<br>Symbol Digit<br>Similarities<br>Go-No-Go |

**Supplementary Table 2.** Normative Sample Characteristics (*N* = 547)

| **Supplementary Table 2. Normative Sample Characteristics (N = 547)** |  |
| --- | --- |
| Age; *avg(SD)* | 65.53 (7.78) |
| Education; *avg(SD)* | 6.95 (4.93) |
| Gender |  |
| % women | 59 |
| Depressive Symptoms |  |
| CESD; *avg(SD)* | 1.85 (1.31) |
| Rurality; % |  |
| 100,000 + | 61.1 |
| 99,999-15,000 | 15.0 |
| 14,999-2,500 | 6.8 |
| <2,500 | 17.2 |
| % Insured | 92 |
| Employment Status |  |
| % Employed | 40.1 |
| % Retired | 18.3 |
| % Unemployed | 41.6 |
| Medical Conditions |  |
| Disease Burden Score; *avg(SD)* | 0.76 (0.82) |
| Hypertension; % | 43.3 |
| Diabetes; % | 23.8 |
| Heart Attack; % | 2.9 |
| Heart Failure; % | 6.2 |
| Stroke; % | 0 |
| CESD = Center for Epidemiologic Studies Depression Scale |  |

**Supplementary Table 3.** Prevalence of MCI with memory Complaints.

| Factor | Any MCI<br>(n = 211) | Amnesic<br>(n = 39) | Language<br>(n = 23) | Visuospatial<br>(n = 54) | Executive<br>(n = 23) | MCDM<br>(n = 43) | MCDN<br>(n = 29) |
| --- | --- | --- | --- | --- | --- | --- | --- |
| Overall | 11.8<br>(10.21-13.39) | 2.2<br>(1.51-2.89) | 1.3<br>(0.77-1.83) | 3.0<br>(2.20-3.80) | 1.3<br>(0.77-1.83) | 2.4<br>(1.68-3.12) | 1.6<br>(1.02-2.18) |
| Age group |  |  |  |  |  |  |  |
| ≤ 66 | 9.5<br>(7.50-11.50) | 2.3<br>(1.29-3.31) | 1.0<br>(0.31-1.69) | 2.2<br>(1.21-3.19) | 1.3<br>(0.53-2.07) | 1.9<br>(0.97-2.83) | 1.5<br>(0.68-2.32) |
| > 66 | 14.4<br>(8.64-20.16) | 2.5<br>(1.38-3.62) | 2.0<br>(0.99-3.01) | 4.5<br>(3.01-5.99) | 1.6<br>(0.69-2.51) | 3.5<br>(2.18-4.82) | 2.1<br>(1.07-3.13) |
| Education |  |  |  |  |  |  |  |
| ≤ 6 years | 13.8<br>(11.74-15.86) | 2.5<br>(1.57-3.43) | 1.7<br>(0.94-2.46) | 4.1<br>(2.92-5.28) | 1.6<br>(0.86-2.34) | 3.1<br>(2.07-4.12) | 2.4<br>(1.49-3.31) |
| > 6 years | 7.2<br>(4.91-9.49) | 2.2<br>(0.90-3.50) | 0.8<br>(0.02-1.58) | 1.6<br>(0.49-2.71) | 1.0<br>(0.12-1.88) | 1.6<br>(0.49-2.71) | 0.4<br>(-0.15-0.95) |
| Sex |  |  |  |  |  |  |  |
| Male | 10.1<br>(7.80-12.40) | 1.3<br>(0.45-2.15) | 1.3<br>(0.45-2.15) | 3.8<br>(2.34-5.26) | 1.5<br>(0.57-2.43) | 2.1<br>(1.00-3.20) | 0.9<br>(0.18-1.62) |
| Female | 13.1<br>(10.91-15.29) | 3.2<br>(2.05-4.35) | 1.5<br>(0.71-2.29) | 3.0<br>(1.89-4.11) | 1.4<br>(0.64-2.16) | 3.1<br>(1.97-4.23) | 2.5<br>(1.48-3.52) |

Note. Values represent % (95% confidence intervals). MCI = mild cognitive impairment; MCDM = multiple domains with memory impairment; MCDN = multiple domains without memory impairment.

**Supplementary Table 4.** Multinomial Logistic Regression Predicting MCI Status with Complaints.

|  | B (SE) | OR | 95% CI for OR |  |
| --- | --- | --- | --- | --- |
|  |  |  | Lower | Upper |
| <u>MCI-Memory vs Unimpaired</u> |  |  |  |  |
| Age | 0.03 (.02) | 1.03* | 1.00 | 1.06 |
| Education | 0.01 (.03) | 1.01 | 0.95 | 1.08 |
| Sex | 0.35 (.23) | 1.42 | 0.80 | 2.51 |
| Rurality | -0.09 (.20) | 0.92 | 0.56 | 1.50 |
| Depressive Symptoms | 0.17 (.04) | 1.18*** | 1.08 | 1.29 |
| Medical Conditions |  |  |  |  |
| Stroke | -0.53 (.48) | 0.59 | 0.08 | 4.46 |
| Hypertension | -0.03 (.20) | 0.97 | 0.60 | 1.56 |
| Diabetes | -0.18 (.20) | 0.84 | 0.49 | 1.45 |
| Heart Disease | -0.59 (.32) | 0.56 | 0.23 | 1.34 |
| Insurance Status | -0.42 (.35) | 0.66 | 0.26 | 1.69 |
| Employment Status |  |  |  |  |
| Retired | 0.19 (.41) | 1.21 | 0.56 | 2.70 |
| Unemployed | 0.46 (.32) | 1.59 | 0.86 | 2.95 |
| <u>MCI-Non-Memory vs Unimpaired</u> |  |  |  |  |
| Age | 0.04 (.01) | 1.04*** | 1.02 | 1.07 |
| Education | -0.06 (.03) | 0.95 | 0.89 | 1.00 |
| Sex | -0.04 (.23) | 0.96 | 0.62 | 1.49 |
| Rurality | 0.16 (.20) | 1.17 | 0.79 | 1.73 |
| Depressive Symptoms | 0.15 (.04) | 1.16*** | 1.08 | 1.25 |
| Medical Conditions |  |  |  |  |
| Stroke | 0.62 (.48) | 1.86 | 0.73 | 4.73 |
| Hypertension | 0.04 (.20) | 1.04 | 0.71 | 1.55 |
| Diabetes | 0.46 (.20) | 1.58* | 1.06 | 2.35 |
| Heart Disease | -0.30 (.32) | 0.74 | 0.39 | 1.39 |
| Insurance Status | -0.27 (.35) | 0.76 | 0.38 | 1.52 |
| Employment Status |  |  |  |  |
| Retired | -0.60 (.34) | 0.55 | 0.28 | 1.07 |
| Unemployed | -0.18 (.23) | 0.84 | 0.53 | 1.32 |

*Note.* Age and education were mean centered, male sex, urban setting, absence of medical condition, being insured, and employed were the reference groups; \*\*\* $p<.001$ , \*\* $p<.01$ , \* $p<.05$ ; MCI=mild cognitive impairment.

